# Cardiovascular Disease Events in Adults with a History of State Care in Childhood: Pooling of Unpublished Results from 9 Cohort Studies

**DOI:** 10.1101/2024.01.26.24301814

**Authors:** G. David Batty, Mika Kivimäki, Ylva B Almquist, Johan G. Eriksson, Mika Gissler, Emmanuel S Gnanamanickam, Mark Hamer, Josephine Jackisch, Hee-Soon Juon, Markus Keski-Säntti, Chaiquan Li, Tuija M. Mikkola, Emily Murray, Amanda Sacker, Leonie Segal, Philipp Frank

## Abstract

**Background:** Individuals who were separated from their biological family and placed into the care of the state during childhood (out-of-home care) are more prone to developing selected physical and mental health problems in adulthood, however, their risk of cardiovascular disease (CVD) is uncertain. Accordingly, we pooled published and unpublished results from cohort studies of childhood care and adult CVD.

**Methods:** We used two approaches to identifying relevant data on childhood care and adult CVD (PROSPERO registration CRD42021254665). First, to locate published studies, we searched PubMed (Medline) until November 2023. Second, with the aim of identifying unpublished studies with the potential to address the present research question, we scrutinised retrieved reviews of the impact of childhood state care on related adult health outcomes. All included studies were required to have prospective measurement of state care in childhood and a follow-up of CVD events in adulthood as the primary outcome (incident coronary heart disease and/or stroke). Collaborating investigators provided study-specific estimates which were aggregated using random-effects meta-analysis. The Newcastle-Ottawa Scale was used to assess individual study quality.

**Findings:** Thirteen studies (2 published, 11 unpublished) met the inclusion criteria, and investigators from nine provided viable results, including updated analyses of the published studies. Studies comprised 611,601 individuals (301,129 women) from the US, UK, Sweden, Finland, and Australia. Relative to the unexposed, individuals with a care placement during childhood had a 50% greater risk of CVD in adulthood (summary rate ratio after basic adjustment [95% confidence interval]: 1.50 [1.22, 1.84]); range of study-specific estimates: 1.28 to 2.06; *I*^2^ = 69%, p = 0.001). This association was attenuated but persisted after multivariable adjustment for socioeconomic status in childhood (8 studies; 1.41 [1.15, 1.72]) and adulthood (9 studies, 1.28 [1.10, 1.50]). There was a suggestion of a stronger state care-CVD association in women.

**Interpretation:** Our findings show that individuals with experience of state care in childhood have a moderately raised risk of CVD in adulthood. For timely prevention, clinicians and policy makers should be aware that people with a care history may need additional attention in risk factor management.

**Research in context:** *Evidence before this study:* There is growing evidence that individuals who were separated from their biological family and placed into the care of the state during childhood (out-of-home care) are more prone to developing selected physical and mental ill-health in adulthood, however, their risk of cardiovascular disease (CVD) events is uncertain. A search of electronic databases to November 2023 yielded only 2 relevant published studies and these had discordant findings.

*Added value of this study:* By scrutinising retrieved reviews of the impact of childhood state care on broadly related adult health outcomes, we identified studies with the potential to examine the association between childhood care and adult CVD events. Investigators from 7 provided these previously unpublished results and, on aggregating them alongside updated analyses from the 2 published studies, we found that, relative to their unexposed peers, adults with experience of state care earlier in life had a 50% greater risk of CVD. There was evidence that this relationship was partially mediating by socioeconomic status in adulthood, and there was a suggestion of a stronger state care–CVD association in women.

*Implications of all the available evidence:* This meta-analysis suggests that, alongside the array of well-document unfavourable social, behavioural, and health outcomes in adulthood, children experiencing state care may additionally have a higher burden of later CVD. For timely prevention, clinicians and policy makers should be aware that people with a care history may need additional attention in risk factor management.

## Introduction

Although decades-long progress in cardiovascular disease (CVD) epidemiology has led to the identification of a series of modifiable risk factors,^1^ their measurement in middle- and older-age populations does not fully explain the occurrence of the condition.^2,3^ This raises the possibility that CVD may have its origins in early life. A series of cohort studies with extended event surveillance have shown that individuals who were overweight, smoked cigarettes, or had higher levels of blood pressure and blood cholesterol in childhood or adolescence were more likely to develop atherosclerotic phenotypes^4–6^ and be diagnosed with CVD^7–14^ in adulthood.

Whereas there is growing evidence implicating these pre-adult physiological and behavioural risk factors in the aetiology of adult CVD, the role of early life psychosocial characteristics is less certain. An increasingly examined exposure in this context is early life adversity. Denoted by an array of characteristics, including maltreatment (e.g., abuse or neglect by family or other trusted adults), parental loss or the threat thereof (e.g., divorce, incarceration), and a stressful home environment (e.g., parental mental illness, addiction),^15^ there is a strong *prima facie* case implicating childhood adversity in the development of adult CVD. That is, relative to unaffected population controls, people experiencing childhood adversity subsequently have a greater prevalence of CVD risk factors, including lifestyle indices such as cigarette smoking, heavy alcohol intake, obesity, and illicit drug use; ^16,17^ are more likely to socioeconomically disadvantaged, as evidenced by higher levels of unemployment, lower occupational prestige, and modest educational attainment;^18^ and have less favourable levels of metabolic, immune, neuroendocrine, and autonomic functioning.^19^

Removal from the biological family into the apparently safer milieu of state care, most commonly in response to significant harm or the risk thereof, represents one of the more severe components of childhood adversity.^15^ As such, an association with later CVD events would be anticipated but has been little-tested. State care – also referred to as out-of-home care, public care, being looked-after, social care, or substitute care – has increased in prevalence in western societies in recent years: while current estimates vary markedly by country and ethnicity, it may be as high as 13%.^20^ In a recent meta-analysis of prospective studies we have shown that adults with a history of state care in childhood experience a doubling in the risk of premature mortality.^20^ While this was partly ascribed to a high occurrence of suicide in adults exposed to early life care,^20^ it is plausible that common chronic diseases in adulthood, specifically CVD, might also contribute. With the two existing studies on pre-adult care and CVD reaching discordant findings,^21,22^ the status of this relationship is uncertain.

The purpose of this systematic review and meta-analysis therefore is to add to the evidence base on early life adversity and adult health by utilising unpublished cohort data on CVD disease rates in individuals with and without a history of state care in childhood. In doing so, we assess if the relationship is confounded by family social circumstances, mediated by adult health behaviour (cigarette smoking) or social status, and whether the care–CVD association varies according to key contexts, including sex, age at care entry, and country.

## Methods

### Search Strategy and Study Selection

We took two approaches to identifying relevant data on childhood care and adult CVD (PROSPERO registration CRD42021254665). First, to locate individual published cohort studies, we searched PubMed. Second, with the aim of identifying unpublished studies with the potential to address the present research question, we scrutinised reviews of the impact of childhood state care on related adult health outcomes.^20,23^ In composing this manuscript, we followed the Meta-analysis Of Observational Studies in Epidemiology (MOOSE) guidelines for content.^24^

The PubMed (Medline) database was searched from its inception in 1966 to November 21, 2023. Without applying any restrictions, we used a series of terms for the exposure (e.g., ‘out-of-home care’, ‘foster care’, ‘public care’, ‘looked after children’) and the outcome (e.g., ‘cardiovascular disease’, ‘coronary heart disease’, ‘stroke’) in the context of longitudinal studies (e.g., ‘cohort’, ‘follow-up’). For a full description of search terms see Supplemental Box 1.

For inclusion, studies needed to satisfy four criteria: a cohort study in which the assessment of care was made prospectively pre-adulthood; focus of the study was out-of-home care and not adoption; data on an unexposed comparator group were available; and a diagnosis of adult CVD events had been made. We then attempted to trace the original study investigators, with wider-ranging internet searches required for authors of older publications. To increase the likelihood of success, multiple authors from the same papers were contacted simultaneously.

### Unpublished Data Sought from Collaborators

After confirmation of the availability of the required data and agreement to participate, an analytical plan was circulated to collaborating investigators. Information sought including analytical sample size, number of people with a care history, number of CVD events, and analysis of the relation between care and adult CVD using either time-to-event analyses or logistic regression as per the available data (Supplemental file, Guidance for Collaborators).

For CVD, defined as comprising incident coronary heart disease and stroke events, three sources of data were regarded as being acceptable. First, registry data for death or hospitalisations from which International Classification of Disease (ICD) codes for coronary heart disease (ICD-9: 410–414; ICD-10: I20– 25) and stroke (ICD-9: 430–438; ICD-10: I60–69) could be extracted. Second, medical examination for coronary heart disease (e.g., electrocardiogram, raised cardiac enzyme activity) and stroke (e.g., computerised tomography scan, magnetic resonance imaging). Third, self-report of a relevant medical condition (e.g., heart attack, myocardial infarction, angina; cerebrovascular disease or accident) or a medical procedure (e.g., coronary artery bypass graft, percutaneous coronary intervention). Self-reported coronary heart disease (kappa statistic 0.70) and stroke (0.66) show good agreement with hospital records.^25^

Where possible, we also requested that investigators adjust for potentially important explanatory factors in their analyses. These included early life socioeconomic status as indexed by parental occupational social class, education, or income, with the substitution of area-based measures if these individual-level data were unavailable. Potential mediating variables requested included the study members’ cigarette smoking habit and socioeconomic status – both captured in adulthood.

### Evaluation of Study Quality

We used the Newcastle-Ottawa Scale to appraise the quality of each study (Supplemental table 1).^26^ For published studies, we assessed existing reports; for unpublished studies we used a combination of publications in which the ascertainment of childhood care or CVD was described plus any other supporting documentation provided by the authors. Comprising eight domains, including the comprehensiveness of exposure and outcome ascertainment and adequacy of the period of health surveillance, a higher score on this scale denotes higher study quality (maximum 9). For the purposes of the present review, studies with a score of 7 or more on the Newcastle-Ottawa Scale were denoted as being of high grade.

### Statistical Analyses

For individual studies using time-to-event analyses, hazard ratios with accompanying 95% confidence intervals were computed using Cox regression.^27^ Where these data were not available, logistic regression was used to calculate odds ratios. In practice, when the health outcome of interest is rare, as is the case in the present analyses for CVD in populations censored by middle-age, odds ratios and hazard ratios will closely approximate. Initially, in the model with basic adjustment, we explore the impact of confounding by controlling for age, sex, or their combination (data from birth cohort studies did not require age adjustment). Family socio-economic status in early life was then added to this model. Next, we examined the role of mediation by social circumstances and cigarette smoking in adulthood. In all these analyses, we observed the change in the risk ratio with basic adjustment after each explanatory variable – confounder or mediator – was added to the multivariable model in a non-accumulative manner.

These study-specific results were pooled using a random effects meta-analysis,^28^ an approach which incorporates the heterogeneity of effects in the computation of their aggregation. An *I*^2^ statistic was computed to summarise this heterogeneity. Lastly, to examine the robustness of our findings, we explored the magnitude of the state care–CVD association (basic adjustment) according to different contexts, including sex, study quality, and geographical region. All analyses were computed using Stata version 17 (StataCorp, College Station, TX), R version 4.3.1, and RStudio version 2023.03.1.

## Results

The search of electronic databases revealed 1 published study matching our inclusion criteria,^21^ while another was published by collaborators during the preparation of this manuscript^22^ (Figure 1). Additionally, we identified eleven unpublished studies from systematic reviews^20^ ^23^ that had the potential to examine the relation between childhood care and adult CVD. In combination, this resulted in 13 unique datasets.^16,19,21,22,29–37^ Requests for collaboration yielded 10 positive responses and 9 study investigators provided viable results which included updated analyses of the two published studies (figure 1 and table 1).

**Figure 1.**
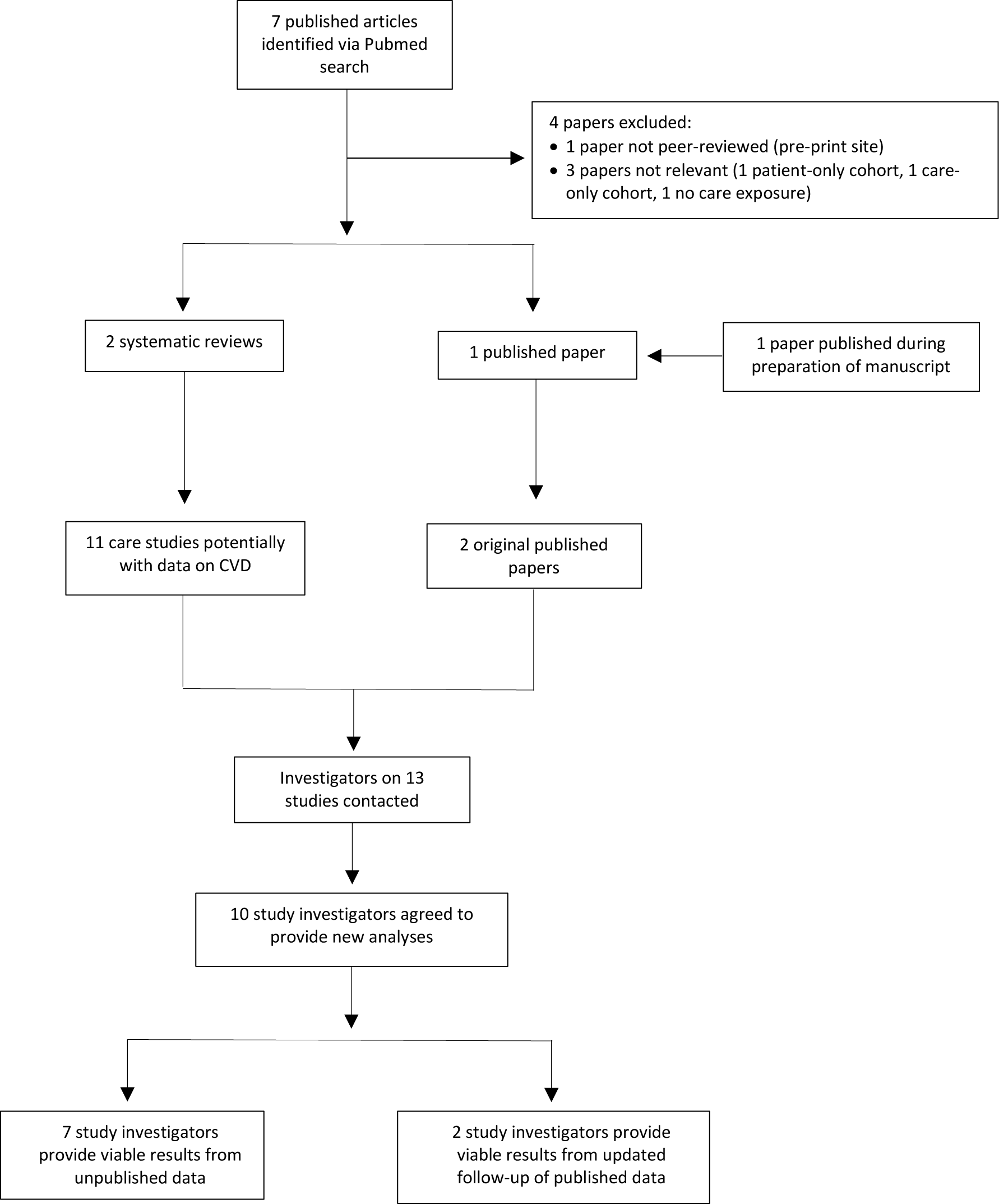
Identification of Published and Unpublished Studies on Childhood Care and Adult CVD.

**Table 1.** Characteristics of Studies Included in the Meta-analysis.

| **Study<sup>key citation</sup> (country)** | **Year of birth** | **Number of participants (women)** | **Age at care assessment (years)** | **Care ascertainment** | **Proportion in care, N (%)** | **Maximum age at follow-up (years)** | **Total number of CVD events** | **CVD ascertainment** |
| --- | --- | --- | --- | --- | --- | --- | --- | --- |
| Helsinki Birth Cohort Study<sup>21</sup> (Finland) | 1934-1944 | 12951 (6119) | 0-11 | Register | 13.3 | 84 | 3578 | Registers of hospitalisations and deaths |
| Stockholm Birth Cohort Study<sup>22</sup> (Sweden) | 1953 | 14543 (7135) | 0-18 | Register | 9.1 | 65 | 1081 | Registers of hospitalisations and deaths |
| Office for National Statistics Longitudinal Study<sup>35</sup> (UK) | 1953-2001 | 353601 (172696) | 1-17 | Carer report | 1.7 | 60 | 503 | Register of deaths |
| 1958 British Birth Cohort Study<sup>16</sup> (UK) | 1958 | 8488 (4371) | 7-16 | Parental report | 3.2 | 58 | 457 | Self-report |
| Woodlawn Cohort Study<sup>33</sup> (USA) | 1960 | 1053 (549) | 6-7 | Parental report | 1.4 | 44 | 34 | Self-report |
| 1970 British Birth Cohort Study<sup>19</sup> (UK) | 1970 | 7888 (4072) | 5-16 | Parental report | 4.7 | 43 | 203 | Self-report |
| impacts of Child Abuse and Neglect (iCAN) South Australia Cohort Study<sup>36</sup> (Australia) | 1972-1999 | 94799 (43851) | 0-18 | Register | 6.6 | 45 | 255 | Registers of hospitalisations, deaths & emergency visits |
| 1987 Finnish Birth Cohort Study<sup>37</sup> (Finland) | 1987 | 59476 (29041) | 0-18 | Register | 3.2 | 33 | 358 | Registers of hospitalisations and deaths |
| 1997 Finnish Birth Cohort Study<sup>31</sup> (Finland) | 1997 | 58802 (28924) | 0-18 | Register | 5.7 | 23 | 66 | Registers of hospitalisations and deaths |

NR, not report. UK, United Kingdom. Studies are ordered by ascending birth year. The iCAN study comprises a Child Protection cohort (record in child protection system from 1986) and a birth cohort (those born from 1986); prior publications have been based on the birth cohort only.

Seven studies were based on samples drawn from Europe^16,19,21,22,31,35,37^ with an additional two from the USA^33^ and Australia^36^ (table 1). In total, these studies comprised 611,601 individuals (301,129 women), with individual cohort size ranging from 1053^33^ to 353,601.^35^ Births occurred across eight decades (1934^21^ to 2001^35^). The period prevalence of state care in childhood varied from 1.4% (USA)^33^ to 13.3% (Finland).^21^ There was a total of 6535 cases of CVD in adulthood; the largest single study recording 3578 such events.^21^ The maximum age at follow-up was 69 years.^21^ Four studies relied on parent/carer reported care history,^16,19,33,35^ while 5 utilised registry data on this exposure.^21,22,31,36,37^ In three cohorts, study members self-reported a physician diagnosis of CVD,^16,19,33^ and in the remaining 6 physician-verified CVD hospitalisations and/or deaths were extracted from national registries.^21,22,31,35–37^ Five of the nine studies were judged as being of higher methodological quality (Supplemental table 1).^21,22,31,35,37^

In Figure 2 we show the study-specific associations between state care ascertained in childhood and subsequent adulthood CVD risk. After the basic adjustments—age alone, sex alone, or a combination—in each of the 9 included studies, the point estimates indicate that a history of state care placement during childhood was related to an elevated risk of adulthood CVD. While these study-specific risk ratios were directionally consistent and above unity, there was clear heterogeneity in their magnitude (range: 1.07 to 2.06, I^2^ 69%, p-value 0.001), and in five studies the care–CVD association was not statistically significant at conventional levels. Aggregating these estimates resulted in around a 50% increase in the risk of CVD in adults with a history of state care in childhood (risk ratio; 95% confidence interval: 1.51; 1.22, 1.86). The Helsinki Birth Cohort differs from the other 8 studies in this meta-analysis in as much as study members entering care did so owing to wartime evacuation to the neighbouring country of Denmark. Excluding results from this study had little impact on the recomputed aggregated estimates (1.59; 1.39, 1.81).

**Figure 2.**
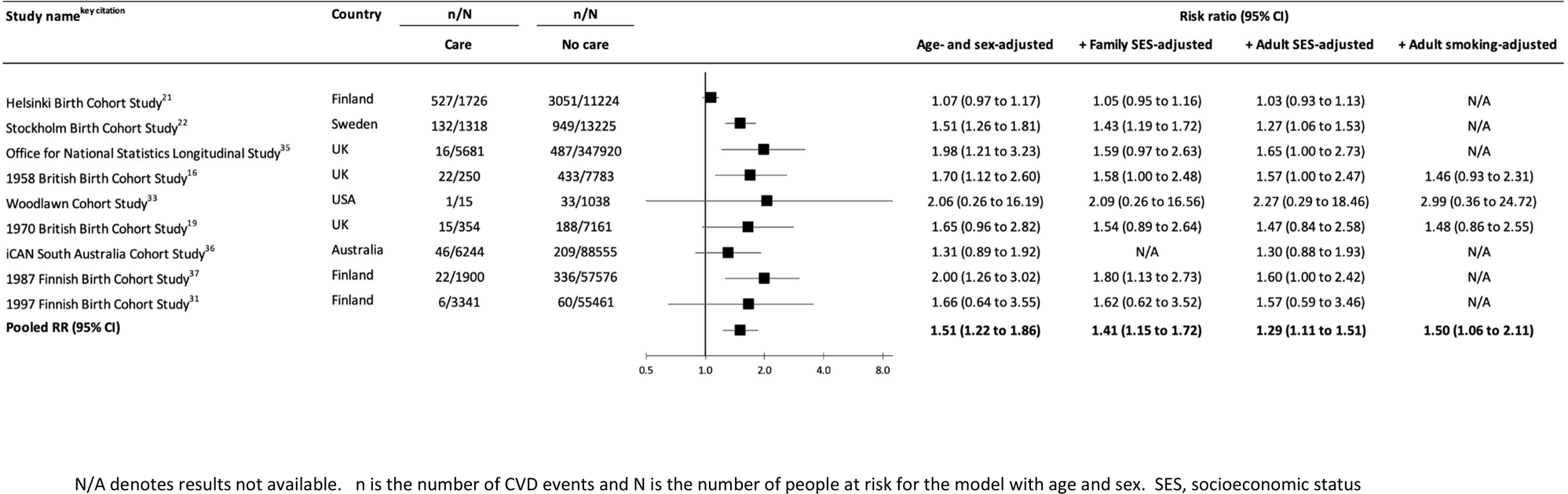
Association Between Public Care in Childhood and Risk of CVD in Adulthood: Meta-analysis of Unpublished Results. N/A denotes results not available. n is the number of CVD events and N is the number of people at risk for the model with age and sex. SES, socioeconomic status

In analyses of the potential role of confounding and mediating factors, there was marginal attenuation of the state care–CVD relation after taking into account childhood (family) social circumstances (8 studies, 1.41; 1.15, 1.72). When study members’ own socioeconomic status in adulthood, a potential mediator, was added to the multivariable model, the relationship between care and CVD was markedly attenuated (9 studies, 1.29; 1.11, 1.51). In contrast, controlling for adult cigarette smoking was not indicative of mediation (1.50; 1.06, 2.11), although this observation was based on only 3 studies with these data.

Results for the impact of different contexts on the care–CVD relation are shown in Figure 3. While there were some differences in the magnitude of the care–CVD association, as evident from the overlapping confidence intervals, these did not differ statistically. Somewhat stronger associations were apparent in women (1.70; 1.29, 2.26) than men (1.29; 1.05, 1.59) and in study participants who were placed in state care later in childhood (1.98; 1.40, 2.79) relative to earlier (1.27; 1.00, 1.61).

**Figure 3.**
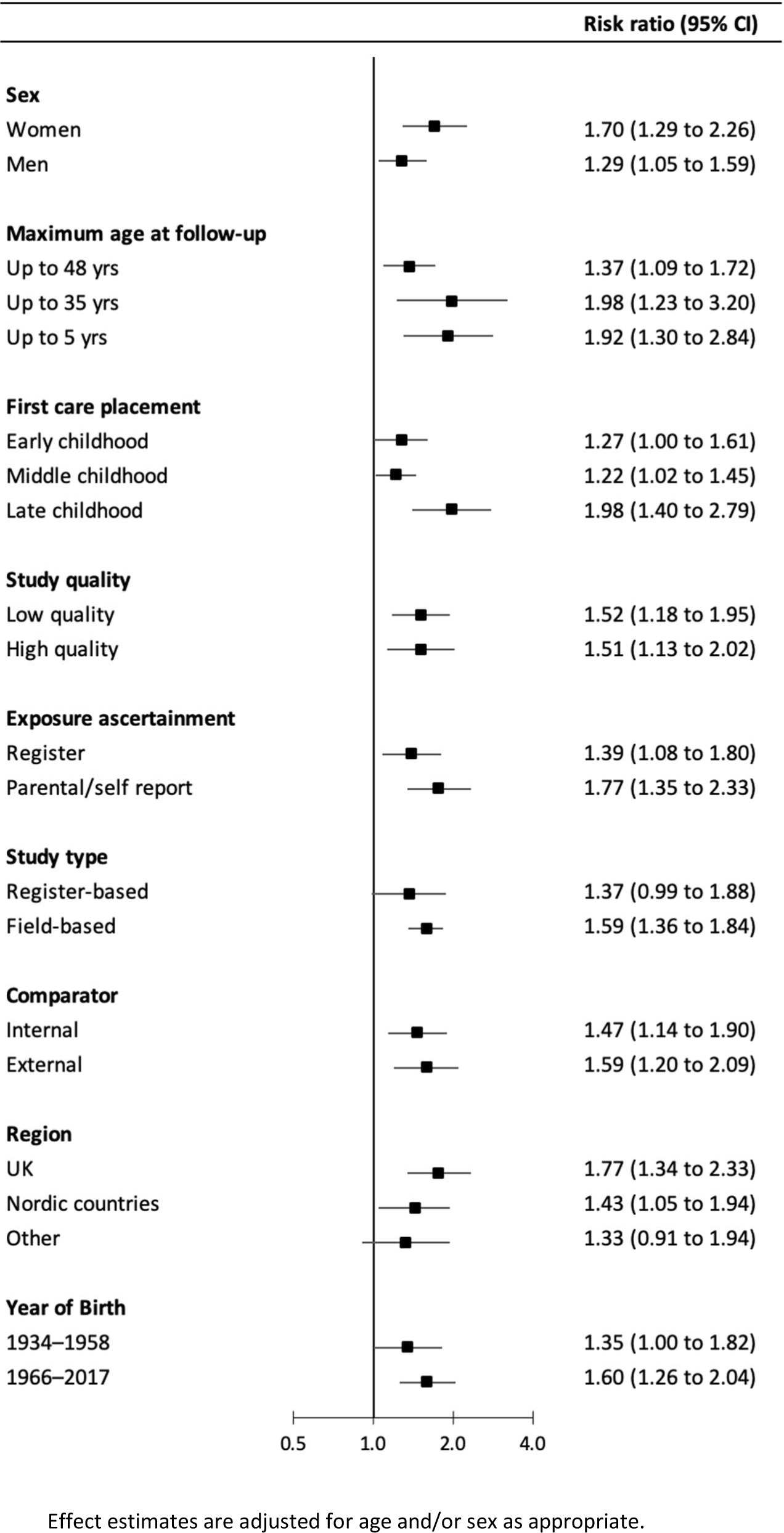
Association Between Public Care in Childhood and Risk of CVD in Adulthood According to Context. Effect estimates are adjusted for age and/or sex as appropriate.

## Discussion

The main finding of this meta-analysis of unpublished results from nine cohort studies was that adults with a history of state care placement in childhood had a moderately raised risk of CVD. The magnitude of this association was commensurate with childhood overweight, cigarette smoking, and raised levels of blood pressure and blood cholesterol.^7–14^ While adjusting for childhood socioeconomic status and adult cigarette smoking had little impact on this association, there was marked attention of effects estimates by social circumstances in later life. When we explored the role of different contexts, there was some suggestion of stronger care—CVD associations in women and individuals entering care later in childhood.

### Comparison with existing studies

Two studies have previously reported on the link between early life care and adult CVD risk and results from these were updated herein.^21,22^ New findings from the Stockholm Birth Cohort Study^22^ were identical to the pooled estimate in the present analyses, while those children in the Helsinki Birth Cohort^21^ who were evacuated from Finland to the ostensibly safer country of Denmark during World War II did not subsequently develop different rates of CVD relative to those remaining with their family of origin (1.07; 0.97, 1.17). The circumstances of this removal from the biological family contrasts with other studies featured in our review, such that Finnish parents volunteered their children for evacuation owing to an abundance of concern for their safety as opposed to them being removed by the state. The pooled estimate following exclusion of this study was not, however, appreciably different to the original. That statistical control for adult social circumstances in our meta-analysis led to a marked attenuation of the care–CVD relationship may indicate that state care places an individual on a trajectory of socioeconomic disadvantage which extends into adult life, an observation made elsewhere when total mortality was the endpoint of interest.^38^

### Effect modification by sex and age at care entry

Our finding of a somewhat stronger relationship of placement in state care with CVD in women than men was also apparent in a recent systematic review in which suicide was the outcome of interest.^20^ These results run counter to speculation that girls are more resilient to stressful early life circumstances than boys.^39^ The observation of sensitive periods of exposure—we found somewhat stronger associations with CVD in people who entered care later in childhood—has been made in relation to other health outcomes, including all-cause mortality, and has an array of plausible explanations.^20^ Older age at care entry could, for instance, simply be a proxy for extended exposure to a dysfunctional home environment. Relatedly, the reasons for care initiation seem to vary by age, such that parental abuse is more common in children entering at younger ages, while behavioural issues (e.g., delinquency) become more prevalent in adolescence.^40^ We attempted to disentangle the impact of pre-care trauma from the effect of care itself by controlling for childhood socioeconomic circumstances. While this had little impact on the magnitude of the care–CVD relationship, the utility of these data for this purpose is low.

### Strengths and Limitations

A strength of our meta-analysis is the focus on studies with childhood care data that were collected prospectively. As exemplified in the much-cited progenitor Adverse Childhood Experiences Study,^41^ and in systematic reviews,^42^ much of what is known about the health impacts of early life adversity, including experience of childhood care, has been gleaned from studies where middle- or older-aged participants responded retrospectively to enquiries about their pre-adult environment.^15^ Based on an aggregation of 20 studies which explored the validity of childhood prospective assessment of maltreatment – the gold standard in these analyses – against distant recall of the same in adulthood, there was low agreement.^43^ This may have important ramifications for studies exploring the links between early adversity and later health endpoints. For instance, prospective measurement of childhood overcrowding revealed no association with adult respiratory disease, whereas when retrospectively-captured, higher levels actually appeared to confer protection against the same outcome.^44^ Similarly discordant were results from a Finnish study in which vascular disease was the outcome of interest.^45^

Our meta-analysis is not without its shortcomings. First, there was some evidence of mediation by adult social circumstances, however, of the health behaviours, we only had data on adult smoking but not physical activity nor alcohol intake. The lack of data on candidate biological mediators, including markers of metabolic, immune, neuroendocrine, and autonomic functioning may not be a limitation, however, given that these characteristics were not related to earlier care exposure in two of the birth cohorts featured here.^19,46^ Second, with the included studies being observational, our results cannot be used to imply cause and effect. An alternative approach to addressing the present question that would circumvent the primary concern of confounding is a randomised controlled trial in which half of children requiring transfer to a safer environment would be allocated to state care while the rest remaining with their family of origin. With such a trial being potentially unethical, a further option is a natural experiment whereby the impact of changes in state care policy (e.g., a new policy to reduce the number of children being placed in out of the home), on CVD risk is explored. Sibling analyses, whereby one child is taken into care but the other remains with the biological family, would also have utility. Third, while we were able to examine the association of age at care entry with CVD, we did not have data on other potentially important care characteristics such as reason for removal to care, care type, and duration across a sufficiently large number of studies to facilitate analyses. That we found marked cross-study differences in care—CVD effect estimates could be ascribed to heterogeneity in exposure such that there is variation in care type across countries (e.g., home versus institution-based). Lastly, with some exceptions,^33,36^ included study samples comprised ethnically white study participants. While it is unlikely that the care–CVD gradient in minority groups would be directionally inconsistent with the present results, empirical testing is warranted.

### Conclusions

Our findings from a pooling of nine cohorts from the US, UK, Sweden, Finland, and Australia show that individuals who experienced state care placement during childhood had a moderately elevated risk of CVD in adulthood. For children with a care history who are therefore known to health and social services, it may be that existing protections are insufficient to address the burden of cardiovascular disease. For timely prevention, clinicians should be aware that children and adults with a care history may need additional attention in risk factor management.

## Supporting information

Guidance for analyses

Systematic review search terms

MOOSE checklist

## Data Availability

All data produced in the present work are contained in the manuscript.

## Contributions

GDB generated the idea for this project, conducted the literature search, contacted study principal investigators, developed an analytical plan, collated results, and drafted the manuscript. PF conducted the meta-analysis, prepared the figures, and edited the manuscript. MK contacted study principal investigators, developed an analytical plan, and edited the manuscript. ESG, MH, JJ, H-SJ, MK-S, CL, TMM and AS analysed individual participant data and edited the manuscript. YBA, JGE, MG, EM, and LS edited the manuscript. All authors were responsible for the decision to submit the manuscript.

## Acknowledgement

LS and ESG acknowledge the South Australia (SA) families and children whose de-identified data contributed to the analyses, the SA-NT DataLink as the data linkage organisation, and the SA Department for Child Protection (DCP) and other SA government agencies that provided study data. AS and EM gratefully acknowledge the permission of the Office for National Statistics to use the Longitudinal Study and the help provided by staff of the Centre for Longitudinal Study Information & User Support (CeLSIUS). CeLSIUS is supported by the ESRC Census of Population Programme (ES/V003488/1). The authors alone are responsible for the interpretation of the data. This work contains statistical data from ONS which is Crown Copyright. The use of the ONS statistical data in this work does not imply the endorsement of the ONS in relation to the interpretation or analysis of the statistical data. This work uses research datasets which may not exactly reproduce National Statistics aggregates. The derivation of 1971 and 1981 NSSEC & Goldthorpe classes is provided in Bukodi and Neuburger (2009) “Data Note. Job and occupational histories for the NSHD 1946 Birth Cohort” as part of the ESRC Gender Network Grant, Project 1 ‘Changing occupational careers of men and women’, Reference: RES-225-25-2001. The code was kindly provided by Erzsebet Bukodi and adapted for use in the LS by Buscha and Sturgis as part of the ESRC grant ‘Inter-cohort Trends in Intergenerational Mobility in England and Wales: income, status, and class (InTIME)’(ES/K003259/1).

## Data sharing

*Bona fide* researchers interested in individual study datasets included in this meta-analysis should contact individual study investigators.

