## Supplementary material for "Cardiovascular Disease Events in Adults with a History of State Care in Childhood: Pooling of Unpublished Results from 9 Cohort Studies": Guidance for analyses

**Guidance for Collaborators**

Thank you for agreeing to participate in this project.

**Study and collaborator details**

| **Study name** |  |
| --- | --- |
| **Country of origin** |  |
| **Details of key supporting paper** with state care as the primary exposure (this will be cited in the manuscript): |  |
| **Authors contributing to this project.** For the purposes of manuscript preparation, for each investigator, please respond to fields opposite. For each additional person, please cut and paste these fields. | Name:  Role (e.g., data analyst, principal/co-investigator):  Salutation (e.g., Professor, Doctor):  Affiliation/s:  Highest research degree:  Email address:  ORCID: |

**Analytical guidance**

Please follow this analytical guidance and populate the following blank tables as extensively as possible. We have tried to keep the requested analyses to a minimum to avoid overburdening collaborators. Any required empirical analyses should be explicit from the tables themselves, footnotes, and other descriptions, but if you have any queries, do of course get in touch.

1. Incident cardiovascular disease (CVD) broadly comprises coronary heart disease and stroke. In table 1, we have described the various CVD data that are of relevance to the present project and their sources. These include International Classification of Disease (ICD) codes based on registry data (death and hospitalisation); self-report of a medical condition or medical procedure; and clinical/study examination.
2. For the primary analyses, our focus is on a single composite CVD outcome: CVD versus no CVD.
3. If you have a sufficiently high number of events for separate analyses of coronary heart disease and stroke in relation to childhood care, these additional results would also be useful. Please replicate table 3 for the reporting of analyses of CHD and/or stroke events where applicable.
4. If you have sufficiently high number of events in men and women, please provide sex-specific results for CVD. This is because there are some reasons to anticipate the impact of care on CVD may differ according to sex.
5. Table 3 may initially appear voluminous, but you may find that some analyses may not be possible in your study. This may be owing to an absence of data (e.g., age at care entry, mediating variables etc), or insufficient numbers of events (e.g., for stroke, or according to sex etc). If so, just leave these cells blank and add a note.
6. To summarise the relationship between care and CVD, we would favour time-to-event analyses. This is most conventionally done using Cox proportional hazards regression to compute hazard ratios with accompanying 95% confidence intervals. Odds ratios, based on logistic regression analyses also with 95% confidence intervals, are less preferable but acceptable if you lack experience in time-to-event analyses. Both techniques will yield very similar results in the context of CVD in the present cohorts.

**Table 1. Guidance for denoting cardiovascular disease, coronary heart disease, and stroke in your study**

| **Disease outcome** | **Source of data** | | |
| --- | --- | --- | --- |
|  | **Registry (death and/or hospitalisation)** | **Medical examination (study- or clinically-based)** | **Self-report (interviewer- or self-administered questionnaire)** |
| Coronary heart disease | ICD-9: 410–414  ICD-10: I20–25 | Electrocardiogram; hospital acute electrocardiogram; raised cardiac enzyme activities. Your study may utilise the MONICA criteria for coronary heart disease. | May include enquiries regarding disease (heart disease, heart attack, myocardial infarction, angina [e.g., Rose questionnaire], peripheral artery disease) or a medical procedure (e.g., coronary artery bypass graft, or percutaneous coronary intervention). |
| Stroke | ICD-9: 430–438  ICD-10: I60–69 | Computerized tomography (CT) scan; Magnetic resonance imaging (MRI) | May include enquiries regarding disease (stroke, cerebrovascular disease/accident) and the sub-types ischaemic stroke (e.g., thrombotic or embolic occlusion of a cerebral artery) and haemorrhagic stroke (e.g., subarachnoid or intraparenchymal haemorrhage). |
| Cardiovascular disease | All the above | All the above | All the above |

**Table 2. Characteristics of your study and its members**

| **Study name** | **Birth years of study members (range)** | **Number of people in analytical sample (number of women)** | **Number of people exposed to childhood care (number of women)** | **Mean (SD) age at baseline (range), years** | **Mean (SD) follow-up (range), years** | **Source(s) of CVD events used in analyses (e.g., deaths, hospitalisations, self-report) and definition (e.g., ICD-codes etc)** |
| --- | --- | --- | --- | --- | --- | --- |

**Table 3. Hazard ratios (95% confidence interval) for the association of childhood care with adult cardiovascular disease outcomes – full cohort (men and women combined)**

| **Statistical adjustment** | **Exposure group** | **Number of people at risk** | **Number of CVD events** | **Hazard/Odds^a^ ratio** | **95% Confidence Interval** |
| --- | --- | --- | --- | --- | --- |
| **Confounder adjustment** |  |  |  |  |  |
| Age- and sex-adjusted (model 1) | No care |  |  | 1.0 (ref) | - |
|  | Care |  |  |  |  |
| Age- and sex-adjusted (model 1) | No care |  |  | 1.0 (ref) |  |
|  | Care started in early childhood^b^ |  |  |  |  |
|  | Care started in middle childhood |  |  |  |  |
|  | Care started in late childhood |  |  |  |  |
| Age-, sex- and early life socioeconomic status-adjusted^c^ (model 2) | No care |  |  | 1.0 (ref) | - |
|  | Care |  |  |  |  |
| **Mediator adjustment** |  |  |  |  |  |
| Model 2 plus subject’s own adult socioeconomic status^d^ | No care |  |  | 1.0 (ref) |  |
|  | Care |  |  |  |  |
| Model 2 plus subject’s own adult smoking habit | No care |  |  | 1.0 (ref) |  |
|  | Care |  |  |  |  |

**^a^**Delete as appropriate

^b^The definitions of early, middle, and late childhood are somewhat arbitrary and based on the Lancet Public Health review (Batty et al. 2022): early childhood (0–6 years), middle childhood (7–12 years); late childhood (13–19 years).

^c^The early life socioeconomic characteristic may include parental occupational social class, parental education, or parental income. An area-based measures can be substituted if these individual-level data are not available. Please control for one factor.

^d^Adult socioeconomic status refers to the study members’ own socioeconomic status later in life – occupational social class, education, or. Again, an area-based measure can be substituted if these individual-level data are not available.

**Table 4. Hazard ratios (95% confidence interval) for the association of childhood care with adult cardiovascular disease outcomes – separate analyses for men and women**

| **Statistical adjustment** | **Exposure group** | **Number of people at risk** | **Number of CVD events** | **Hazard/Odds^a^ ratio** | **95% Confidence Interval** |
| --- | --- | --- | --- | --- | --- |
| **Men** |  |  |  |  |  |
| Age-adjusted | No care |  |  | 1.0 (ref) | - |
|  | Care |  |  |  |  |
| Age-, and early life socioeconomic status^b^-adjusted | No care |  |  | 1.0 (ref) | - |
|  | Care |  |  |  |  |
| **Women** |  |  |  |  |  |
| Age-adjusted | No care |  |  | 1.0 (ref) | - |
|  | Care |  |  |  |  |
| Age- and early life socioeconomic status-adjusted | No care |  |  | 1.0 (ref) | - |
|  | Care |  |  |  |  |

**^a^**Delete as appropriate

^b^The early life socioeconomic characteristic may include parental occupational social class, parental education, or parental income. An area-based measures can be substituted if these individual-level data are not available. Please control for one factor.
