## Supplementary material for "Cardiovascular Disease Events in Adults with a History of State Care in Childhood: Pooling of Unpublished Results from 9 Cohort Studies": Systematic review search terms

**Supplemental Box 1. Search string for PubMed (Medline)**

#1 (cardiovascular disease[Title/Abstract]) OR (coronary artery disease[Title/Abstract]) OR (coronary heart disease[Title/Abstract]) OR (myocardial infarction[Title/Abstract]) OR (ischemic heart disease[Title/Abstract]) OR (ischaemic heart disease[Title/Abstract]) OR (acute coronary syndrome[Title/Abstract]) OR (stroke[Title/Abstract]) OR (cerebrovascular accident[Title/Abstract]) OR (cerebrovascular disease[Title/Abstract]) OR (cardiovascular events[Title/Abstract]) OR (cardiovascular deaths[Title/Abstract]) OR (heart failure[Title/Abstract])

#2 (out-of-home care [Title/Abstract]) OR (out of home care [Title/Abstract]) OR (foster care [Title/Abstract]) OR (public care [Title/Abstract]) OR (looked-after-children [Title/Abstract]) OR (looked after children[Title/Abstract])

#3 (epidemiologic studies[MeSH Terms]) OR (cohort studies[MeSH Terms]) OR (epidemiologic[Text Word]) OR (longitudinal[Text Word]) OR (cohort[Text Word]) OR (follow up[Text Word]) OR (observational[Text Word]) OR (prospective[Text Word])

#4 #1 AND #2 AND #3

**Supplemental Table 1. Cohort study quality assessment according to the**

**Newcastle-Ottawa criteria: Meta-analysis**

| **Study name^key citation^** | **Selection** | | | | **Comparability** | **Outcome** | | | **Total quality score (0-9)** |
| --- | --- | --- | --- | --- | --- | --- | --- | --- | --- |
|  | **Representative(0-1)** | **Selection – unexposed**  **(0-1)** | **Ascertainment exposure (0-1)** | **Outcome absent at baseline (0-1)** | **Case/control comparability**  **(0-2)** | **Assessment of outcome (0-1)** | **Length of follow-up**  **(0-1)** | **Adequacy of follow-up**  **(0-1)** |  |
| Helsinki Birth Cohort Study^1^ | 0 | 1 | 1 | 1 | 2 | 1 | 1 | 1 | 8 |
| Stockholm Birth Cohort Study^2^ | 0 | 1 | 1 | 1 | 2 | 1 | 1 | 1 | 8 |
| Office for National Statistics Longitudinal Study^3^ | 1 | 1 | 1 | 1 | 2 | 1 | 1 | 0 | 8 |
| 1958 British Birth Cohort Study^4^ | 1 | 1 | 0 | 1 | 1 | 0 | 1 | 1 | 5 |
| Woodlawn Cohort Study^5^ | 1 | 1 | 0 | 1 | 1 | 0 | 1 | 0 | 5 |
| 1970 British Birth Cohort Study^6^ | 1 | 1 | 0 | 1 | 1 | 0 | 1 | 0 | 5 |
| iCAN South Australia Cohort Study^7^ | 0 | 1 | 1 | 1 | 2 | 1 | 0 | 0 | 6 |
| 1987 Finnish Birth Cohort Study^8^ | 1 | 1 | 1 | 1 | 2 | 1 | 0 | 0 | 7 |
| 1997 Finnish Birth Cohort Study^9^ | 1 | 1 | 1 | 1 | 2 | 1 | 0 | 0 | 7 |

A higher score denotes higher study quality
